# Multivariate Machine Learning Analysis of M-ECG-derived Heart Rate Variability in TBI Veterans, With and Without Current PTSD and Additional Psychiatric Comorbidity

**DOI:** 10.64898/2026.06.05.26354915

**Authors:** Aqil Izadysadr, Hamideh Sadat Bagherzadeh, Jared A. Rowland, Sarah L. Martindale, Jennifer R. Stapleton-Kotloski, Dwayne W. Godwin

## Abstract

Traumatic brain injury (TBI) and posttraumatic stress disorder (PTSD) frequently co-occur in Veterans, producing overlapping symptoms and shared autonomic dysregulation. Heart rate variability (HRV) offers a noninvasive measure of autonomic function. This study explored whether multivariate HRV features extracted from MEG-derived electrocardiogram (M-ECG) signals contained information related to group classification of Veterans with TBI alone (TBI-alone; n = 42) versus those with current PTSD, with or without additional psychiatric comorbidity (TBI+PTSD; n = 40). Time-domain, frequency-domain, geometric, and nonlinear HRV metrics were analyzed using a nested cross-validated Random Forest classifier, with Boruta-based feature selection and SHapley Additive exPlanations for model interpretability. The classifier demonstrated modest exploratory discrimination (Random Forest AUC = 0.663, 95% CI: 0.540-0.778). LF/HF ratio, low-frequency proportion, and approximate entropy were among the features contributing most to distinguishing the TBI+PTSD group, suggesting that multivariate HRV analysis of M-ECG signals may capture subtle, yet potentially informative, autonomic patterns associated with current PTSD and psychiatric burden among Veterans with TBI. The modest classification performance indicates that these findings should be considered preliminary and hypothesis-generating. Larger, independent studies are needed to determine the robustness and potential clinical relevance of these findings.

## 1. Introduction

Traumatic brain injury (TBI) is a major cause of long-term neurological and autonomic dysfunction (Esterov and Greenwald, 2017) and is common among Veterans seeking clinical care (Brenner et al., 2013; Hoffman and Taylor, 2019; Walker et al., 2025). Individuals with TBI may experience affective symptoms, altered stress responsivity, cardiovascular dysregulation, and cognitive and sensorimotor impairments (Prathep et al., 2014; Logan et al., 2018; Hoffman and Taylor, 2019; Howlett et al., 2022). Posttraumatic stress disorder (PTSD) is one of the most common and clinically consequential comorbidities following TBI (Wall, 2012; Kaplan et al., 2018; Van Praag et al., 2019). There exists substantial overlap in symptom presentation in comorbid TBI and PTSD (Kaplan et al., 2018). Both TBI and PTSD have been independently associated with autonomic nervous system dysregulation, characterized by altered autonomic regulation (Esterov and Greenwald, 2017; Pertab et al., 2018; Schneider and Schwerdtfeger, 2020). Veterans, in particular, show a significantly higher rate of co-occurring PTSD and TBI than the general civilian population (Loignon et al., 2020), which makes them a valuable cohort for evaluating autonomic dysregulation.

Heart rate variability (HRV) provides a noninvasive measure of autonomic nervous system function by quantifying variation in the time intervals between successive heartbeats. It encompasses multiple dimensions, including time-domain measures of vagal tone, spectral indices of autonomic regulation, and nonlinear metrics that characterize regulatory complexity. Optimal HRV has been linked to better health, greater self-regulatory capacity, and increased adaptability or resilience (Shaffer and Ginsberg, 2017). For instance, higher resting levels of vagally mediated HRV have been linked to better executive functioning, including attentional control and emotional processing supported by the prefrontal cortex (McCraty and Shaffer, 2015). Conversely, diminished variability reflects age-related decline, chronic stress, disease processes, or weakened regulation across multiple levels of control (Singer et al., 1988; Malik et al., 1996; Thayer et al., 2009).

Prior work connects PTSD to altered HRV; meta-analytic findings demonstrate reduced root mean square of successive differences (RMSSD), high-frequency power, and overall variability indexed by the standard deviation of NN intervals (SDNN), together with an elevated resting heart rate; higher LF/HF ratios were also observed in unadjusted analyses, suggesting diminished parasympathetic modulation and altered baseline autonomic regulation in individuals with PTSD (Schneider and Schwerdtfeger, 2020).

Autonomic dysfunction has also been documented in TBI and is marked by reduced HRV across mild, moderate, and severe injuries, with abnormalities that may persist into sub-acute and chronic stages. The most affected metrics in TBI include reduced time-domain measures, including mean RR interval, SDNN, and RMSSD, and decreased absolute high-frequency and total spectral power (Talbert et al., 2024). A population-based study of adults with a history of mild TBI demonstrated that in mild TBI, persistent autonomic dysfunction may affect vagally mediated time-domain indices, including RMSSD and percentage of NN intervals that differ by more than 50 ms (Bat-Erdene et al., 2026). Greater HRV reductions in TBI are generally associated with increasing injury severity, and lower HRV reliably predicts poorer recovery, worse functional outcomes, and increased mortality following TBI (Talbert et al., 2024). These findings indicate the clinical relevance of autonomic dysregulation in TBI and demonstrate its potential role as a physiological marker of injury-related and recovery-related processes (Pinto et al., 2024). They also raise the possibility that PTSD comorbidity introduces additional or distinct patterns of autonomic disruption within TBI populations. Evidence from cross-sectional research indicates that reduced HRV is significantly associated with the combined presence of pain, PTSD, and mild TBI among combat Veterans (Tan et al., 2009; Ellis and Watanabe, 2025). In addition, LF power has been reported to increase in individuals with co-occurring TBI and PTSD (Weaver et al., 2018; Cowansage et al., 2025).

While there is growing interest in HRV as a physiological measure of autonomic regulation, HRV measures are highly interrelated and reflect different but overlapping aspects of autonomic regulation; therefore, reliance on individual measures may provide an incomplete characterization of autonomic differences (Kleiger et al., 2005). This limitation is especially relevant in populations such as Veterans with TBI and PTSD, where overlapping symptomatology and physiological heterogeneity complicate group differentiation.

Multivariate machine learning approaches offer an exploratory framework for addressing these challenges. In TBI research, machine learning applied to HRV has shown promise for identifying physiologically and clinically relevant patterns associated with injury severity (Kuruwita Arachchige et al., 2025). HRV-derived features combined with machine learning have also shown promise for predicting survival outcomes in severe TBI (Zhang et al., 2020).

Similarly, in PTSD, machine learning approaches using HRV have been explored to classify diagnostic status (Reinertsen et al., 2017; Sadeghi et al., 2020). However, to date, no study has applied machine learning to identify combinations of HRV features that differentiate individuals with TBI alone from those with TBI and current PTSD, with or without additional psychiatric comorbidity, in Veteran populations.

Recent advances in magnetoencephalography (MEG) have enabled the extraction of cardiac signals directly from MEG data (Godwin et al., 2024; Izadysadr et al., 2026b). MEG-derived electrocardiogram (M-ECG) yields physiologically valid HRV measures comparable to those obtained from a conventional electrocardiogram. This approach allows HRV to be examined retrospectively in existing MEG datasets, where a concurrent electrocardiogram recording was not acquired (Izadysadr et al., 2026b).

Given the heterogeneity of autonomic patterns in TBI and PTSD and the limited research on multivariate HRV signatures in comorbid populations, this study used M-ECG to take an exploratory, data-driven approach to examine HRV patterns across time-, frequency-, geometric, and nonlinear domains that may contain group-related information regarding Veterans with TBI alone versus those with current PTSD, with or without additional psychiatric comorbidity.

## 2. Methods

### 2.1. Data

This retrospective study used data previously collected as part of the Chronic Effects of Neurotrauma Consortium Study 34 (Rowland et al., 2024). The dataset uses the Clinician-Administered PTSD Scale for DSM-5 (CAPS-5; Weathers et al., 2018) to determine the presence or absence of lifetime and current PTSD (0 = absent and 1 = present) as well as the Mid-Atlantic

Mental Illness Research, Education, and Clinical Center Assessment of Traumatic Brain Injury (Rowland et al., 2020) to determine the presence/absence of lifetime TBI history (0 = absent and 1 = present) (Rowland et al., 2024). The Structured Clinical Interview for DSM diagnoses was administered to determine the presence of psychiatric disorders (0 = no current diagnosis and 1 = at least one current diagnosis). The study received approval from the Atrium Health Wake Forest Baptist institutional review board (IRB00036287).

Participants were included if they had a lifetime history of mild TBI (TBI = 1). Participants were classified into the TBI group without current PTSD (TBI-alone) if they had no current PTSD diagnosis, no lifetime history of PTSD, and no current psychiatric disorder (current PTSD = 0; lifetime PTSD = 0; current psychiatric disorder = 0). Participants were classified into the TBI group with current PTSD (TBI+PTSD) if they met criteria for current PTSD. Because PTSD is classified as a current psychiatric disorder, all participants in this group also met the criterion for a current psychiatric disorder (current psychiatric disorder = 1). Consequently, the TBI+PTSD group included individuals with PTSD both with and without additional current psychiatric comorbidities. Participants who did not meet the criteria for either group were excluded from the study. The final sample consisted of 42 participants in the TBI-alone group and 40 participants in the TBI+PTSD group.

Magnetoencephalography data were acquired using a whole-head CTF Systems Inc. MEG 2005 neuromagnetometer equipped with 275 first-order axial gradiometer coils. Resting-state recordings were obtained while participants were seated upright and instructed to sit quietly with eyes open for 5 minutes. MEG signals were sampled at 1200 Hz with a DC-300 Hz acquisition bandwidth and preprocessed using synthetic third-order gradient balancing, whole-trial DC offset correction, and band-pass filtering from DC to 80 Hz, including a 60 Hz notch filter (Rowland et al., 2024).

The study pipeline is shown in Figure 1. Third-party libraries used in data preprocessing include NumPy, Pandas, SciPy, MNE-Python, Matplotlib, AntroPy, and Nolds, which supported numerical computation, neurophysiological signal processing, visualization, and nonlinear time-series analysis (Hunter, 2007; McKinney, 2010; Gramfort et al., 2013; Schölzel, 2019; Harris et al., 2020; Virtanen et al., 2020; Larson et al., 2024; The pandas development team, 2024; Vallat, 2025). Additional libraries for machine learning and statistical analysis included Statsmodels, Boruta, Scikit-learn, SHAP, and Seaborn (Kursa and Rudnicki, 2010; Seabold and Perktold, 2010; Pedregosa et al., 2011; Lundberg and Lee, 2017; Waskom, 2021). All computations were performed in Python, and the custom analytical scripts of this study, including full documentation to facilitate replication, are made available on GitHub at https://github.com/izadysadr/mecg-hrv-ml-tbi-ptsd.

**Figure 1.**
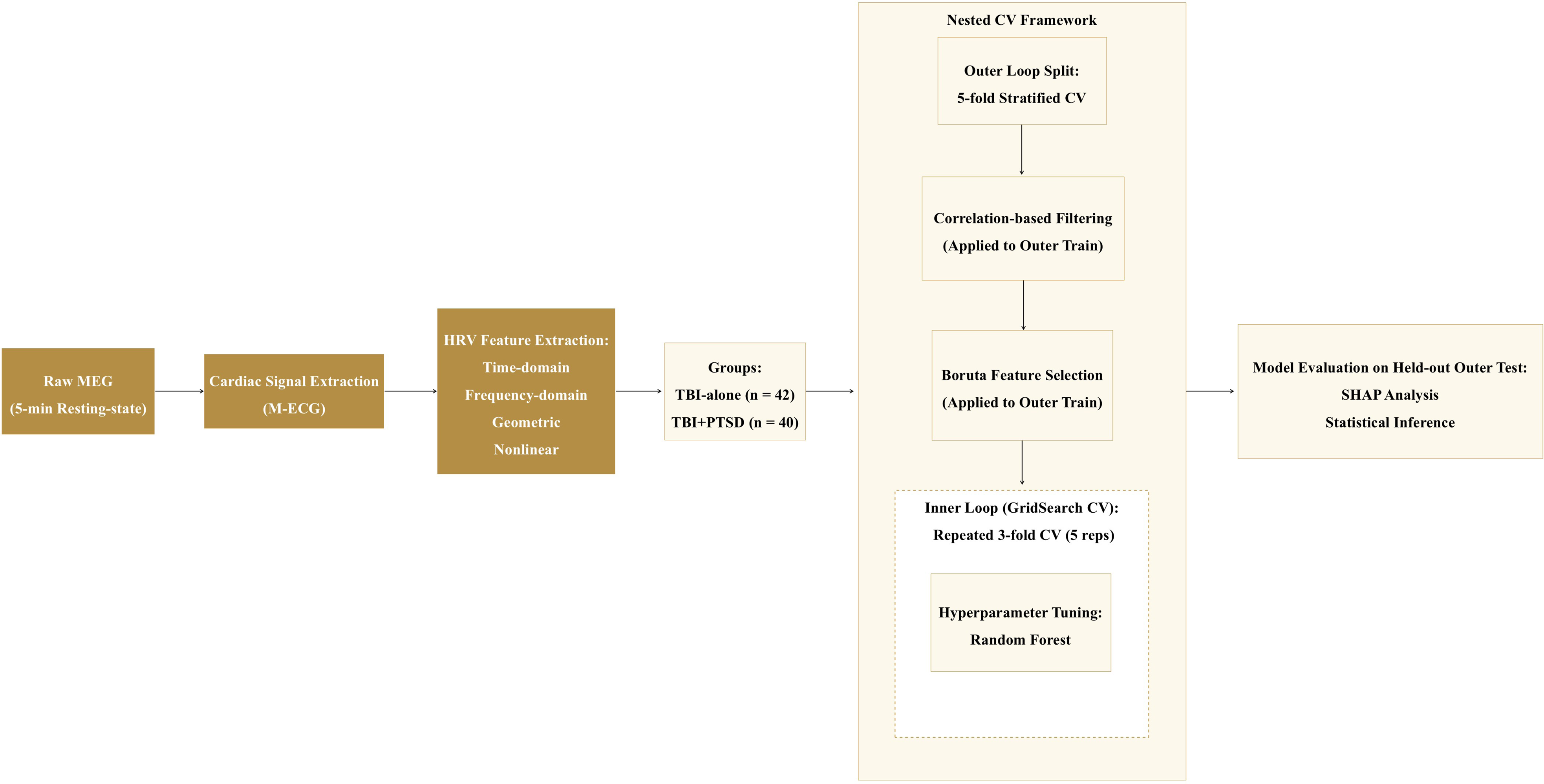
Overview of the study pipeline, illustrating the extraction of heart rate variability (HRV) metrics from MEG-derived electrocardiogram (M-ECG), the nested cross-validation (CV) modeling framework, and subsequent model interpretability analyses.

### 2.2. Preprocessing

M-ECG signals were extracted from the MEG data using procedures described by Izadysadr et al. (2026). Following M-ECG extraction, custom Python scripts were used to compute 18 HRV metrics from the complete 5-minute recording, resulting in one feature vector per participant. The metrics spanned time-, frequency-, geometric-, and nonlinear domains (Table 1). This allowed for characterization of various aspects of autonomic regulation while minimizing redundancy and preserving physiological interpretability. The custom scripts implement an extensive HRV extraction pipeline beyond the scope of the present manuscript; therefore, the complete implementation, documentation, and source code are provided in the accompanying GitHub repository to facilitate reproducibility. Although very-low-frequency (0.0033-0.04 Hz) power was included among the extracted features for completeness, it was not interpreted, given that reliable physiological interpretation of the very-low-frequency power band typically requires longer recording durations.

**Table 1.**
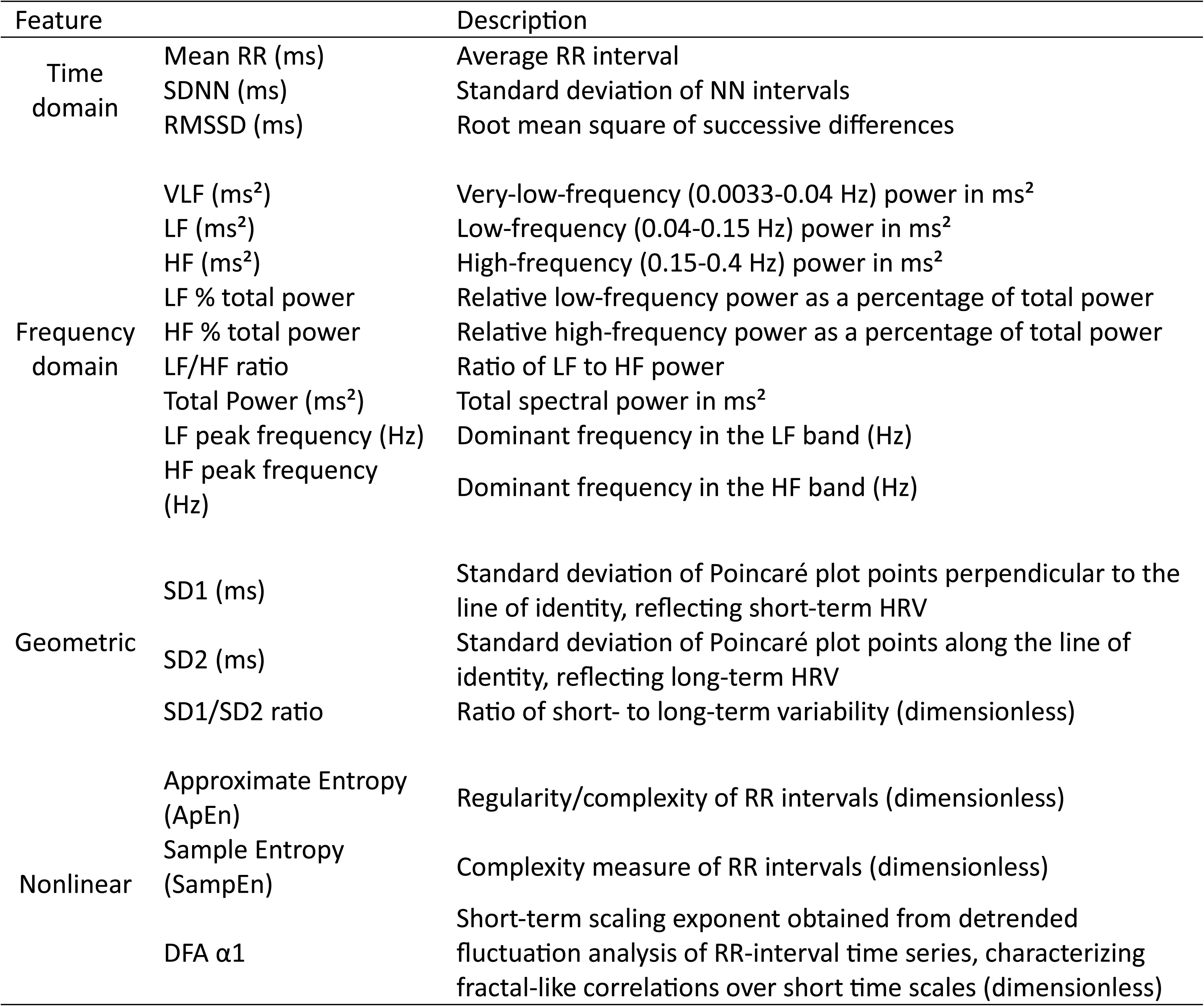
Heart rate variability (HRV) features extracted from 5-min resting-state MEG-derived electrocardiogram (M-ECG) signals. Time-domain features quantify overall and short-term variability of RR intervals. Frequency-domain features include absolute and relative spectral power, peak frequencies, and sympathovagal balance. Geometric features are derived from Poincaré plots, and nonlinear features measure the complexity and fractal characteristics of heart rate dynamics. Units are provided for each feature where applicable.

### 2.3. Machine learning modeling framework

#### 2.3.1. Feature selection

The full feature matrix (X) consisted of 82 subjects and 18 HRV features (82 × 18), while the binary outcome variable (*y*) was a binary target indicating class membership (coded as 0 = TBI-alone and 1 = TBI+PTSD). All 18 HRV features listed in Table 1 were included in the feature matrix, with no prior feature exclusion.

#### 2.3.2. Cross-validation and correlation-based filtering

A nested cross-validation (CV) strategy was implemented to ensure unbiased performance estimation and hyperparameter optimization. This was performed at the participant level, such that each participant contributed a single observation and appeared in exactly one outer test fold across the five-fold CV procedure. The outer loop used a 5-fold stratified CV, preserving class balance across folds, and was used exclusively for model evaluation. Within each outer training set, a repeated stratified 3-fold CV (5 repetitions; total of 15 resampling iterations) was applied for hyperparameter tuning. All CV procedures used shuffling with a fixed random seed (42) to ensure reproducibility. Supplemental Table 1 details the parameters used in the machine learning modeling framework for this study.

For each outer fold, the total number of samples and the number of samples per class were explicitly tracked for both training and test sets. This ensured that stratification was preserved and that no fold exhibited pathological class imbalance that could bias performance estimates.

Missing data were assessed for all HRV features prior to modeling. No missing values were observed in any feature (0% missingness). Consequently, no imputation or missing-data handling was required, and all models were trained on the original feature values.

A correlation-based filtering step was applied within each outer training fold to reduce multicollinearity and eliminate redundant predictors. Pairwise Spearman rank correlation coefficients were computed among all HRV features in the training set, and the absolute correlation matrix was examined. Features with pairwise absolute Spearman correlation > 0.90 (based on the upper triangle of the correlation matrix) were removed. No minimum feature-retention threshold was imposed; all remaining non-redundant features after correlation filtering were passed to the subsequent Boruta procedure. The resulting list of dropped features was derived exclusively from the training data and then applied identically to the corresponding test fold. The frequency with which each feature was removed across folds was recorded to evaluate the stability and redundancy of HRV features across different data partitions.

#### 2.3.3. Wrapper-based feature selection using Boruta

Following correlation-based filtering, wrapper-based feature selection was performed using the Boruta algorithm (Kursa and Rudnicki, 2010) within each outer training fold to identify features whose importance exceeded that of randomized shadow features. Because Boruta performs all-relevant feature selection by comparing observed feature importance against randomized shadow features, it is well-suited for HRV data characterized by nonlinear relationships, correlated predictors, and a limited number of biologically meaningful features where false-negative exclusions are undesirable.

Boruta was implemented using a Random Forest classifier (Breiman, 2001) with the number of trees automatically determined, a maximum tree depth of 10, a fixed random seed of 42, and parallel computation enabled across all available central processing unit cores. The procedure was run for up to 50 iterations with a significance level of α = 0.10 and automatic determination of the number of shadow features. Both confirmed important and tentatively important (“weak”) features were conservatively retained to minimize false-negative exclusion of potentially informative physiological markers, whereas features classified as rejected by Boruta were excluded from model training.

#### 2.3.4. Model selection and training

A tree-based ensemble classifier, namely Random Forest, was evaluated for HRV-based classification because it is well suited to physiological feature sets characterized by nonlinear relationships, correlated predictors, interaction effects, noise, and limited sample sizes.

The classifier was trained using hyperparameters optimized via grid search within the inner CV loop, with ROC AUC (area under the receiver operating characteristic curve) as the selection metric. The hyperparameters included the number of trees (100, 200), maximum tree depth (3, 5), minimum samples required to split a node (5, 10), minimum samples required at a leaf node (2, 5), and a square-root feature subsampling strategy.

No feature scaling or normalization was applied prior to model training, as tree-based methods are invariant to monotonic feature transformations; this also preserved interpretability of feature attributions, namely SHapley Additive exPlanations (SHAP) values, with respect to the original HRV feature scales. HRV feature distributions were inspected to confirm the absence of extreme outliers that could destabilize tree-based splits. Hyperparameter ranges were deliberately constrained to limit model complexity and reduce overfitting, with all tuning performed exclusively within the inner CV loop.

#### 2.3.5. Model evaluation

Final model evaluation was conducted exclusively on the outer test folds of the nested CV procedure. For each fold and each model, performance was assessed using ROC AUC, accuracy, precision, recall (sensitivity), and F1-score, with safeguards applied to precision, recall, and F1-score calculations to prevent division-by-zero errors. To characterize classification error structure, confusion matrices were computed using predicted class labels from the outer test folds. Predictions were then concatenated across all outer folds to construct pooled confusion matrices for each model, providing a single, aggregate estimate of classification errors across all held-out samples and avoiding biases introduced by averaging fold-level confusion matrices. A pooled confusion matrix was computed from the raw prediction counts, from which sensitivity, specificity, precision, and F1-score were derived to summarize overall classification performance.

Model interpretability was assessed using SHAP (Lundberg and Lee, 2017) for the classifier to provide transparent, model-consistent explanations of how individual HRV features contributed to predictions. SHAP was selected because it offers a theoretically grounded, additive feature attribution framework based on Shapley values, enabling both local (sample-level) and global (dataset-level) interpretation while remaining well-suited to complex, nonlinear tree-based models.

A tree-based SHAP explainer was fitted for the Random Forest classifier, and SHAP values were computed exclusively on the held-out test set of each outer CV fold to prevent information leakage. For each fold, sample-level SHAP values and the corresponding test-set feature matrices were retained and pooled across folds to enable global aggregation of feature contributions at the sample level.

Because correlation-based filtering and Boruta feature selection yielded different feature subsets across folds, SHAP values were aligned to a common feature space defined as the union of all selected features, with features not selected in a given fold assigned zero contribution.

Test-set feature matrices were aligned in the same manner to ensure correspondence between SHAP values and feature values. Using this aligned representation, global interpretability was assessed via SHAP summary visualizations, including summary plots to characterize the distribution and directionality of feature effects and dependence plots for the top-ranked features to examine relationships between feature magnitude and model contribution across the pooled test data.

#### 2.3.6. Leakage prevention

Reproducibility and leakage prevention were ensured by fixing random seeds for all stochastic components. Every preprocessing step, including correlation-based filtering, feature selection, and hyperparameter optimization, was performed strictly on the training data within each fold and only then applied to the corresponding held-out test data. At no stage were test data used during model selection or feature engineering.

### 2.4. Statistical analysis

#### 2.4.1. Performance uncertainty estimation

To quantify uncertainty in classification performance, a nonparametric bootstrap was applied to pooled predictions from the outer test folds. Predicted class labels and probabilities derived exclusively from held-out outer-fold test sets were aggregated across folds and treated as a single evaluation set containing predictions from all 82 participants. From this pooled evaluation set, 10,000 bootstrap resamples were generated, each containing 82 observations sampled with replacement. For each resample, accuracy, precision, recall, and F1-score were computed using predicted class labels, and ROC AUC was computed using predicted probabilities. Precision, recall, and F1-score incorporated explicit zero-division handling, assigning zero in degenerate resamples lacking positive predictions or positive true labels, and resamples containing only a single outcome class were excluded from ROC AUC computation. For each metric, the bootstrap distribution was summarized by its mean and a 95% confidence interval defined by the 2.5^th^ and 97.5^th^ percentiles. A fixed random seed (42) was used to ensure reproducibility.

#### 2.4.2. SHAP-based global feature importance

To summarize feature importance at the population level, pooled sample-level SHAP values were transformed to their absolute magnitudes, capturing the strength of each feature’s contribution irrespective of direction. Global importance was then quantified using non-parametric bootstrapping (n = 10,000) over the pooled SHAP distributions, with the mean absolute SHAP value computed for each feature in each bootstrap iteration. Feature-wise importance estimates were reported as the bootstrap mean together with 95% confidence intervals derived from the 2.5^th^ and 97.5^th^ percentiles of the bootstrap distributions.

## Results

### 3.1. CV and Boruta

The TBI-alone group (n = 42) had a mean age of 42.8 ± 10.6 years and was predominantly male, with 38 (90.5%) participants coded as male and 4 (9.5%) coded as female. The TBI+PTSD group (n = 40) had a mean age of 39.7 ± 9.2 years and was similarly predominantly male, with 35 (87.5%) participants coded as male and 5 (12.5%) coded as female. There were no significant differences between groups in age (Mann-Whitney U test, U = 681.5, p = 0.142) or sex distribution (Fisher’s exact test, p = 0.735).

Stratified sampling preserved class balance in all folds, with training sets of 65-66 participants and test sets of 16-17 participants, and no fold exhibited pathological class imbalance. Correlation-based filtering removed a consistent set of highly redundant HRV features across folds, including absolute high-frequency power, total power, and Poincaré plot measures (SD1 and SD2), reflecting intercorrelations among absolute power and geometric metrics. This stable pattern indicates effective reduction of multicollinearity prior to Boruta-based feature selection (Table 2).

**Table 2.**
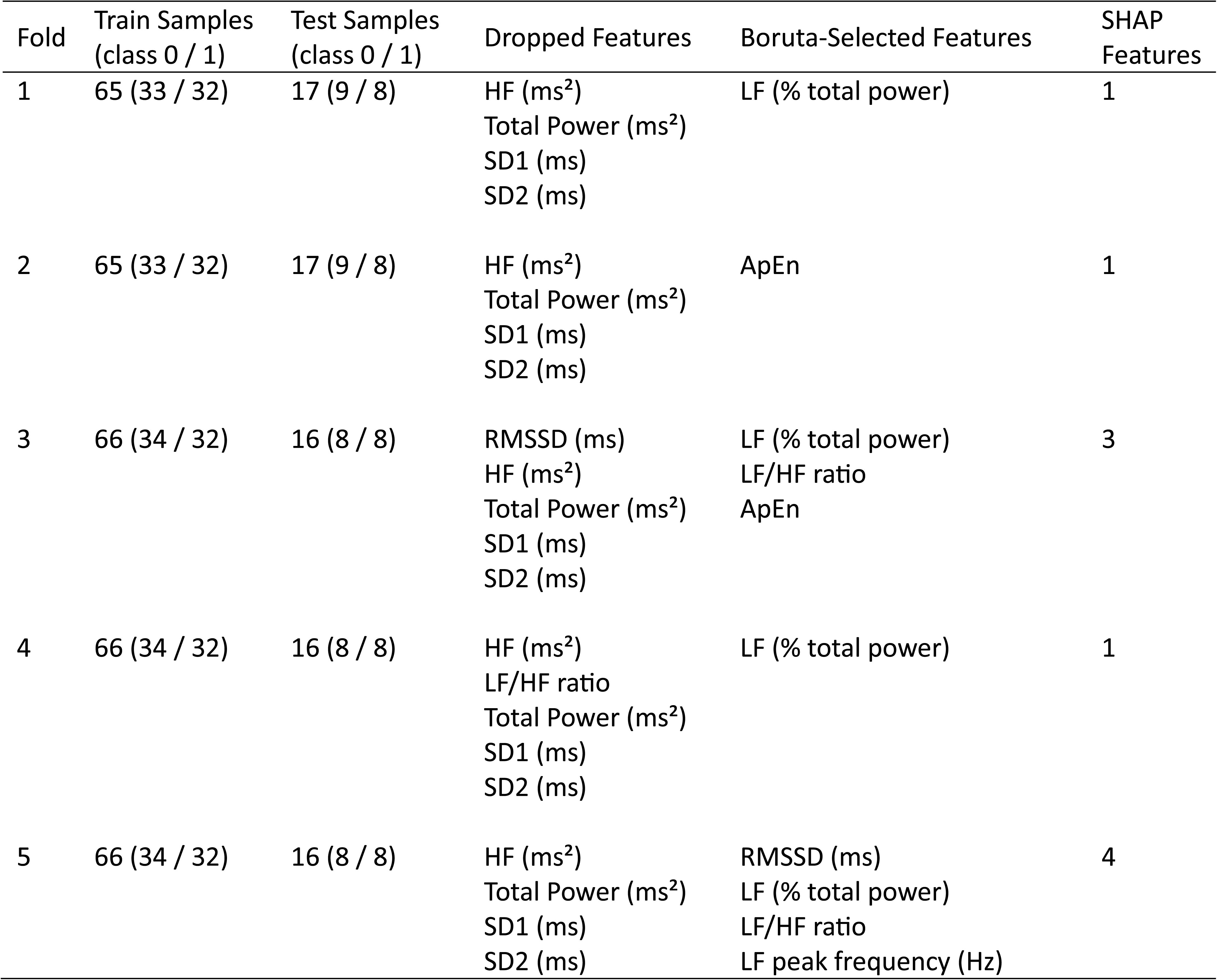
Training and test sample counts, dropped features, Boruta-selected features, and the number of features used for SHapley Additive exPlanations (SHAP) analysis across 5 cross-validation (CV) folds (class 0 = TBI-alone; class 1 = TBI+PTSD). Abbreviations: root mean square of successive differences (RMSSD), relative low-frequency power as a percentage of total power (LF % total power), high-frequency power (HF), dominant frequency in the low-frequency band (LF peak frequency), total spectral power (Total Power), ratio of low-to high-frequency power (LF/HF ratio), standard deviation of Poincaré plot points perpendicular to the line of identity, reflecting short-term HRV (SD1), standard deviation of Poincaré plot points along the line of identity, reflecting long-term HRV (SD2), and approximate entropy, reflecting regularity/complexity of RR intervals (ApEn). Units are provided for each feature where applicable.

Boruta selection yielded compact feature subsets ranging from one to four features per fold for subsequent SHAP analysis. Retained features included relative low-frequency power (LF % total power), approximate entropy (ApEn), LF/HF ratio, RMSSD, and LF peak frequency, spanning time-, frequency-, and nonlinear HRV domains. No fold yielded an empty feature set (Table 2).

### 3.2. Model performance

The ROC curve generated from pooled out-of-fold predictions across the 5-fold outer CV indicated modest discriminative ability above chance level (AUC = 0.663; Figure 2; Table 3).

**Figure 2.**
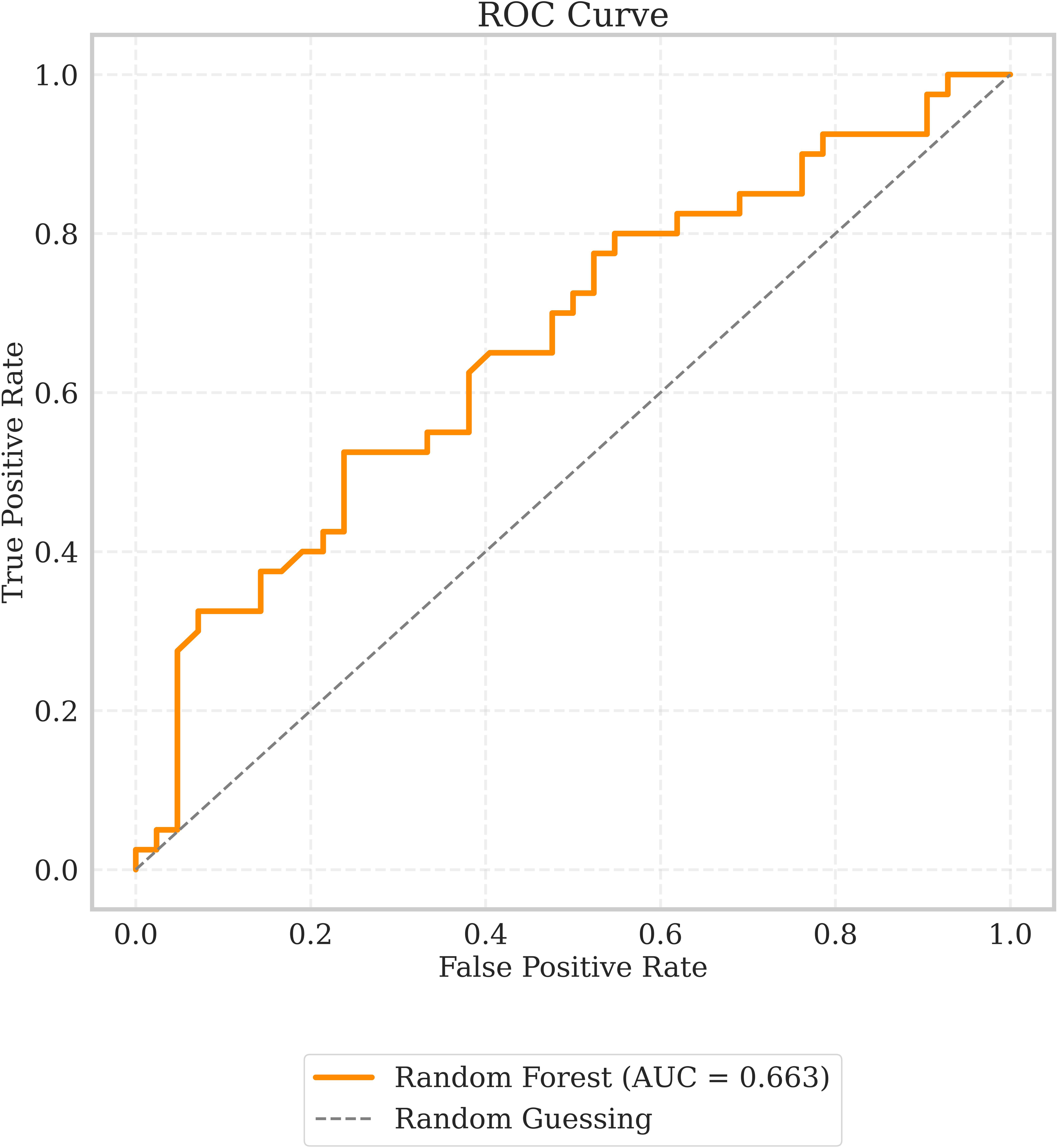
Receiver operating characteristic (ROC) curves based on pooled predictions from the 5-fold outer cross-validation (CV), illustrating model discrimination performance.

**Table 3.**
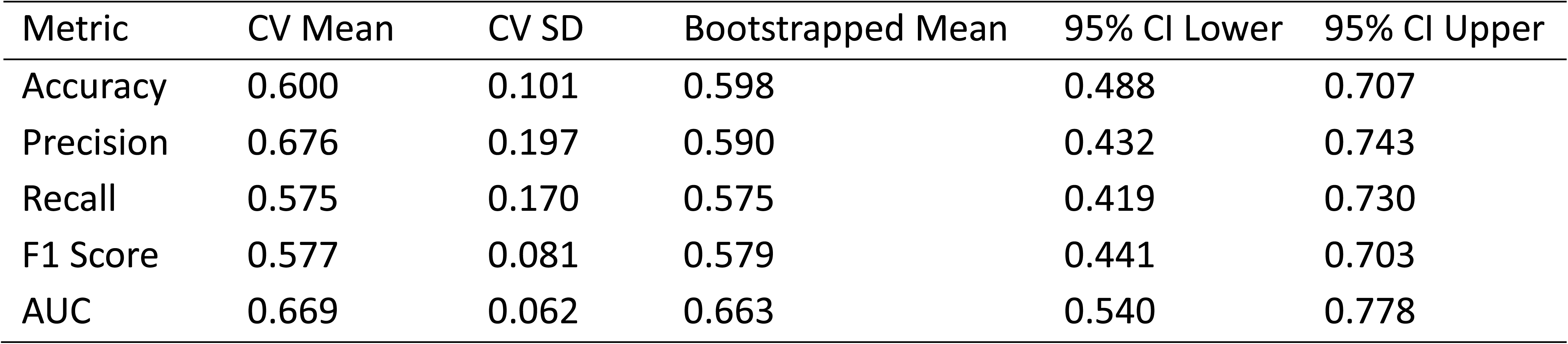
Cross-validated and bootstrapped performance of the Random Forest classifier. Five-fold cross-validation (CV) results are reported as mean ± standard deviation (SD). Bootstrap estimates were obtained from 10,000 resamples of the test dataset and are reported as mean values with 95% confidence intervals (CI). Metrics include accuracy, precision, recall, F1 score, and area under the receiver operating characteristic curve (AUC).

Evaluation across the 5-fold CV yielded a mean AUC of 0.669 ± 0.062 and an accuracy of 0.600 ± 0.101. Nonparametric bootstrapping (n = 10,000) estimated mean performance and 95% confidence intervals. Mean accuracy was 0.598 (95% CI: 0.488-0.707), with precision of 0.590 (95% CI: 0.432-0.743) and recall of 0.575 (95% CI: 0.419-0.730), resulting in an F1 score of 0.579 (95% CI: 0.441-0.703). The corresponding AUC was 0.663 (95% CI: 0.540-0.778) (Table 3).

Pooled confusion-matrix results showed broadly similar classification performance across the two classes. The F1 score was slightly higher for TBI-alone than TBI+PTSD (0.612 vs 0.582), with recall of 0.619 and 0.575, respectively, whereas specificity was higher for TBI+PTSD than TBI-alone (0.619 vs 0.575) (Table 4).

**Table 4.** Summary of pooled confusion matrix metrics for the Random Forest (RF) classifier across all test folds. Metrics include true positives (TP), false positives (FP), false negatives (FN), true negatives (TN), recall (sensitivity), specificity, precision, and F1 score for each class. Values are calculated from the concatenated predictions across folds, providing an overall assessment of model performance for each class (0 = TBI-alone and 1 = TBI+PTSD)

| Model | Class | TP | FP | FN | TN | Recall (sensitivity) | Specificity | Precision | F1 Score |
| --- | --- | --- | --- | --- | --- | --- | --- | --- | --- |
| RF | 0 | 26 | 17 | 16 | 23 | 0.619 | 0.575 | 0.605 | 0.612 |
|  | 1 | 23 | 16 | 17 | 26 | 0.575 | 0.619 | 0.590 | 0.582 |

### 3.3. SHAP

The SHAP summary (violin) plot illustrates the distribution and directionality of feature contributions to model predictions across the pooled outer CV test samples (Figure 3). Based on mean absolute SHAP values, LF % total power had the largest contribution to model predictions, followed by ApEn, LF/HF ratio, LF peak frequency, and RMSSD, indicating that these features contributed the most to the model’s discrimination between TBI-alone and TBI+PTSD groups.

**Figure 3.**
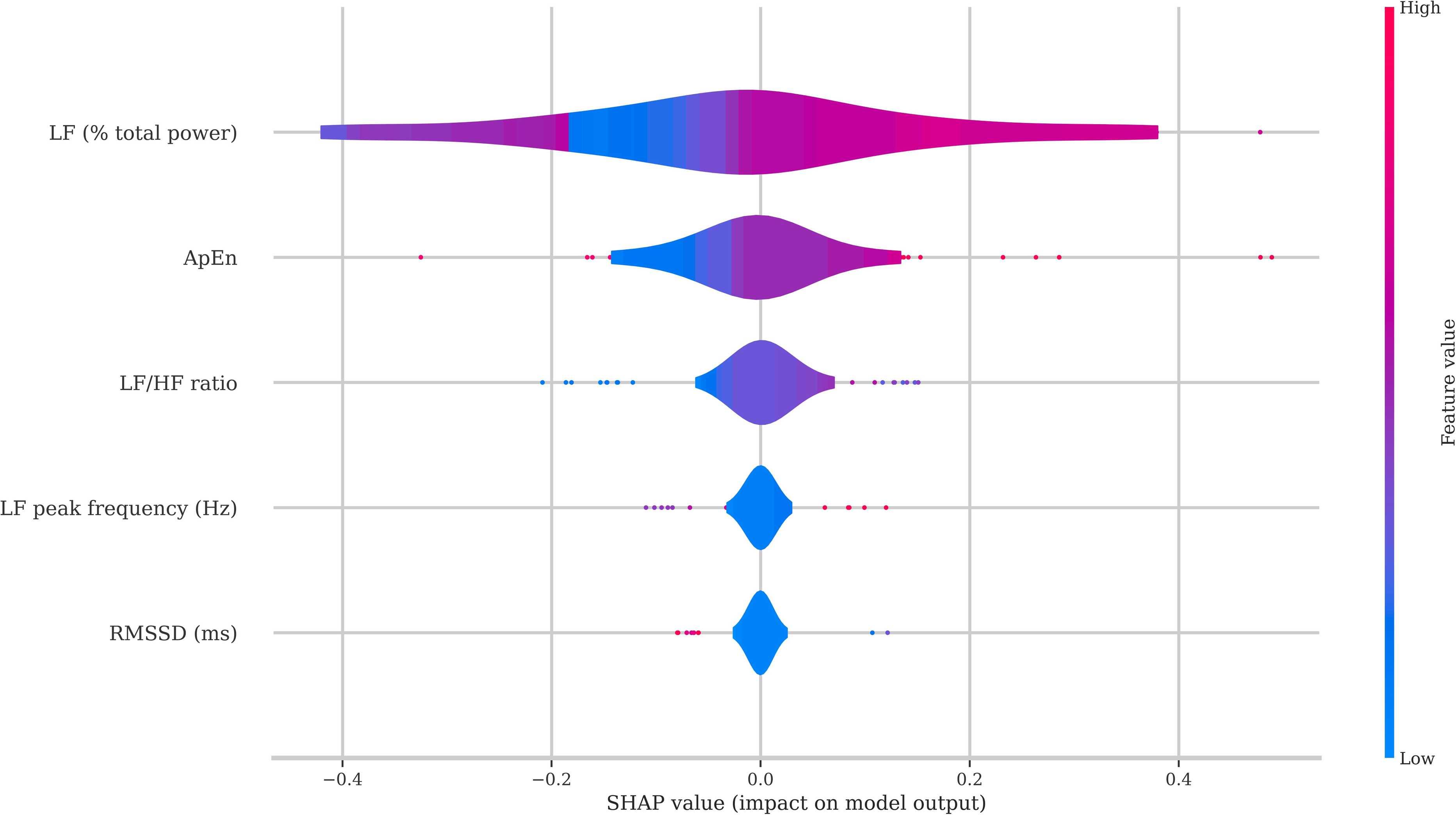
SHapley Additive exPlanations (SHAP) summary violin plot illustrating the distribution and directionality of feature contributions to model predictions across the pooled outer cross-validation (CV) test samples.

SHAP dependence plots for the three highest-ranked features based on mean absolute SHAP values showed distinct patterns of feature influence on model predictions (Supplemental Figure 1). LF % total power exhibited a non-linear relationship with the model output, with intermediate values associated with a higher predicted probability of TBI+PTSD, whereas lower and higher values were associated with a lower predicted probability of TBI+PTSD. Higher ApEn values and higher LF/HF values were also associated with higher predicted probabilities of TBI+PTSD.

Pooled mean absolute SHAP values were further summarized using nonparametric bootstrapping (n = 10,000). LF % total power showed the largest mean absolute SHAP value (0.129, 95% CI: 0.100–0.158), followed by ApEn (0.057, 95% CI: 0.037–0.080) and the LF/HF ratio (0.035, 95% CI: 0.023–0.049). LF peak frequency (0.013, 95% CI: 0.007–0.020) and RMSSD (0.010, 95% CI: 0.005–0.016) showed smaller contributions to model output (Table 5). A mean absolute SHAP value of 0.129 indicates that LF % total power contributed, on average, an absolute magnitude of approximately 0.129 to the model’s predicted probability.

**Table 5.** Bootstrapped SHapley Additive exPlanations (SHAP) feature importance for heart rate variability (HRV) features. Absolute SHAP values, expressed as probabilities, were pooled across all test sets, and 95% confidence intervals (CI) were estimated via bootstrap resampling (n = 10,000). Feature definitions: average RR interval (Mean RR); root mean square of successive differences (RMSSD); relative low-frequency power as a percentage of total power (LF % total power); ratio of LF to HF power (LF/HF ratio); approximate entropy (ApEn).

| Feature | Mean SHAP | 95% CI Lower | 95% CI Upper |
| --- | --- | --- | --- |
| LF (% total power) | 0.1287 | 0.1002 | 0.1581 |
| ApEn | 0.0573 | 0.0367 | 0.0802 |
| LF/HF ratio | 0.0352 | 0.0228 | 0.0486 |
| LF peak frequency (Hz) | 0.0133 | 0.0068 | 0.0203 |
| RMSSD (ms) | 0.0102 | 0.0051 | 0.0160 |

### 3.4. Post-hoc multicollinearity and sensitivity assessment

Computed variance inflation factors for the HRV features retained by Boruta indicated that all selected features, including LF % total power, LF/HF ratio, RMSSD, LF peak frequency, and ApEn, had low variance inflation factor values (range: 1.21-2.29), well below known thresholds for concern (variance inflation factor > 5 or 10; O’Brien, 2007), indicating minimal redundancy among predictors (Table 6).

**Table 6.** Variance inflation factors (VIFs) for the reduced heart rate variability (HRV) feature set. VIF values quantify the degree of multicollinearity among predictors, with all retained features exhibiting low multicollinearity (*VIF* < **33**), indicating that redundancy among features was minimal prior to model training and interpretation. Feature definitions: relative low-frequency power as a percentage of total power (LF % total power); ratio of low-frequency to high-frequency power (LF/HF ratio); root mean square of successive differences (RMSSD); dominant frequency in the low-frequency band (LF peak frequency); approximate entropy, reflecting regularity/complexity of RR intervals (ApEn).

| Feature | VIF |
| --- | --- |
| LF (% total power) | 2.2912 |
| LF/HF ratio | 2.1871 |
| RMSSD (ms) | 1.3109 |
| LF peak frequency (Hz) | 1.2307 |
| ApEn | 1.2075 |

Including sex or age as predictors did not meaningfully affect model performance or feature selection. Bootstrapped AUCs remained comparable to the primary analysis (sex: 0.691 vs 0.663; age: 0.655 vs 0.663). Sex was never selected (0/5 folds), while age was selected in only 2/5 folds and had lower SHAP importance than the principal HRV features (LF % total power, ApEn, and LF/HF ratio). This suggests that neither demographic variable primarily drove model predictions.

## 4. Discussion

In this study, we applied a multivariate machine learning framework to HRV data to test whether HRV features contained information related to group status. The TBI+PTSD group included participants with current PTSD both with and without additional psychiatric comorbidities, whereas the TBI-alone group excluded current psychiatric disorders. This means that the classifier should be interpreted as distinguishing Veterans with TBI and PTSD-related psychiatric burden from those with TBI without current psychiatric morbidity, rather than identifying PTSD-specific autonomic signatures. Although an analysis restricted to isolated PTSD with comorbid TBI would have been informative, only 11 participants met these criteria, precluding a reliable nested CV machine learning analysis.

The Random Forest classifier achieved modest above-chance discrimination with a mean ROC AUC of 0.669 across outer CV folds and a pooled bootstrap AUC of 0.663 (95% CI: 0.540–0.778). The inclusion of sex or age as covariates did not meaningfully alter performance, suggesting that demographic adjustment did not substantially change the observed classification results.

Across folds, LF % total power was selected in all five folds, while ApEn and LF/HF ratio were selected in two folds. The remaining features were selected less consistently. This suggests some instability in feature selection, which may partly reflect the modest sample size and the resulting differences in training data across folds.

SHAP analyses revealed that higher LF/HF ratio and ApEn were associated with an increased probability of TBI+PTSD classification, whereas LF % total power exhibited a non-linear effect where intermediate values predicted TBI+PTSD classification the most. Alterations in LF power have previously been reported in individuals with TBI+PTSD (Weaver et al., 2018; Cowansage et al., 2025).

These findings suggest that, relative to TBI-alone, TBI+PTSD individuals may exhibit subtle differences across a few HRV features, potentially reflecting differences in autonomic regulation, characterized by altered sympathovagal balance (higher LF/HF ratio) (Nelushi et al., 2025) and greater complexity or reduced temporal regularity (higher ApEn) (Shaffer and Ginsberg, 2017). LF power has been linked to baroreflex modulation (Moak et al., 2007; Goldstein et al., 2011; Rahman et al., 2011). The finding that intermediate LF % total power values characterize the TBI+PTSD group may point to altered baroreflex-related autonomic modulation compared to the TBI-alone cohort. One plausible interpretation is that rather than simply indicating higher basal sympathetic tone, this shift may reflect altered baroreflex function or a heightened autonomic responsiveness introduced by PTSD in the context of TBI.

The use of M-ECG to extract HRV demonstrates the feasibility of deriving physiologically valid cardiac signals from MEG datasets, expanding opportunities for retrospective autonomic analyses in studies where conventional ECG recordings are unavailable. Although the observed HRV patterns may have potential relevance to autonomic characterization in the studied cohort, the modest classification performance does not support clinical application at this stage.

In this study, the sample size was modest, and participants were predominantly male, reflecting the veteran population from which the sample was drawn. The TBI+PTSD group had a greater psychiatric burden, and the observed autonomic signatures may reflect the combined influence of PTSD and commonly co-occurring psychiatric conditions.

Because a healthy control group was not included, absolute deviations in HRV and the independent contributions of TBI versus PTSD could not be determined; therefore, our findings should be interpreted as relative differences between the TBI+PTSD and TBI-alone groups.

While nested CV and bootstrapping provide uncertainty estimates of model performance, external validation in independent cohorts is needed to confirm reproducibility. This study focused on mechanistic HRV signatures, and including clinical symptom measures was outside the scope of the current work. In addition, medication information, cardiovascular status, and time-of-scan factors known to transiently influence HRV, including immediate prior caffeine, nicotine, or alcohol consumption, acute pain levels, respiration rate, and sleep quality the night before the scan, were not available for analysis.

Future studies could explore the influence of medications on HRV and assess the incremental value of HRV features over symptom measures. Integrating HRV features with other physiological and neuroimaging markers could improve classification performance and enhance mechanistic understanding. Longitudinal studies could evaluate whether multivariate HRV signatures predict functional outcomes or response to interventions in TBI and PTSD populations. Future work could also examine inter-individual variability in autonomic regulation after TBI, as HRV alterations may emerge only in a subset of individuals. Finally, prospective studies are needed to determine whether pre-existing autonomic dysregulation may represent a vulnerability factor for PTSD.

## 5. Conclusion

This study suggests that multivariate machine learning applied to HRV features derived from M-ECG may may contain information related to group classification of Veterans with TBI alone versus those with current PTSD, with or without additional psychiatric comorbidity. The modest classification performance indicates that these findings should be considered preliminary and hypothesis-generating. Larger, independent studies are needed to determine the robustness and potential clinical relevance of these findings.

## Data Availability

This study retrospectively used data previously collected as part of the Chronic Effects of Neurotrauma Consortium Study 34 (Rowland et al., 2024). The derived analyses are included in the article and/or supplementary material. The analysis code is publicly available on GitHub (https://github.com/izadysadr/mecg-hrv-ml-tbi-ptsd). Further inquiries can be directed to the corresponding authors.

## Nomenclature (List of Abbreviations)

ROC AUC: area under the receiver operating characteristic curve
ApEn: approximate entropy
CV: cross-validation
HF: high-frequency power
HRV: heart rate variability
LF: low-frequency power
LF/HF ratio: ratio of low-frequency to high-frequency power
M-ECG: MEG-derived electrocardiogram
MEG: magnetoencephalography
PTSD: posttraumatic stress disorder
RMSSD: root mean square of successive differences
SDNN: standard deviation of NN intervals
SHAP: SHapley Additive exPlanations
TBI: traumatic brain injury

## Acknowledgments

This study was previously made available as a preprint on medRxiv under the title “Multivariate Machine Learning Analysis of M-ECG-derived Heart Rate Variability in TBI Veterans, With and Without Comorbid PTSD” by Izadysadr et al.

## Ethics statement

The studies involving humans were approved by the Atrium Health Wake Forest Baptist Institutional Review Board (IRB00036287). This study involved analysis of previously collected datasets and was conducted in accordance with local legislation and institutional requirements. Written informed consent was not required in accordance with national legislation and institutional requirements.

## Conflict of Interest Statement

The authors declare that the research was conducted in the absence of any commercial or financial relationships that could be construed as a potential conflict of interest.

## Funding

The author(s) declare that financial support was received for the research, authorship, and/or publication of this article. This work was supported by the Department of Defense Chronic Effects of Neurotrauma Consortium (CENC) Award W81XWH-13-2-0095 and the Department of Veterans Affairs CENC Award I01 CX001135. This work was also supported by grant AA016852 to DG and by the resources of the Salisbury VA Healthcare System. AI, HB, and JS-K were supported by the Department of Neurology.

## Notes

### Competing Interest Statement

The authors have declared no competing interest.

### Author Declarations

The study received approval from the Atrium Health Wake Forest Baptist institutional review board (IRB00036287).

### Summary of Updates

The manuscript has been revised for concision, and structural changes have been made to the supplemental material, table, and figure.

