## Supplemental File for "Multivariate Machine Learning Analysis of M-ECG-derived Heart Rate Variability in TBI Veterans, With and Without Current PTSD and Additional Psychiatric Comorbidity"

**Supplemental Figures and Tables**

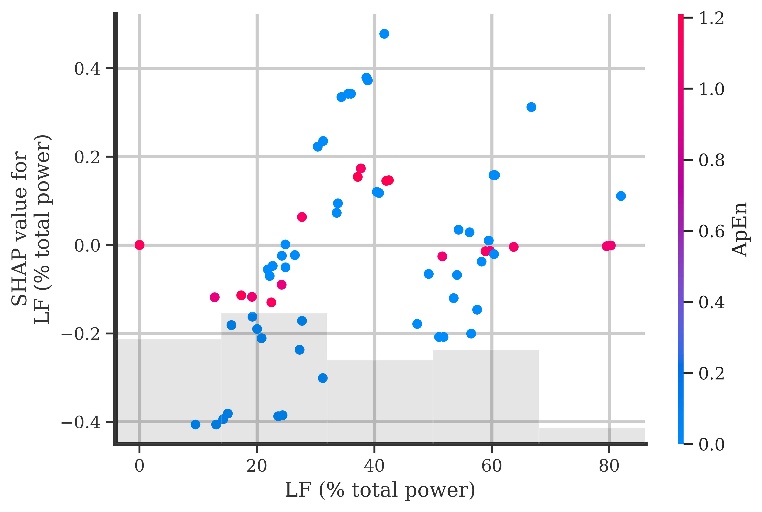

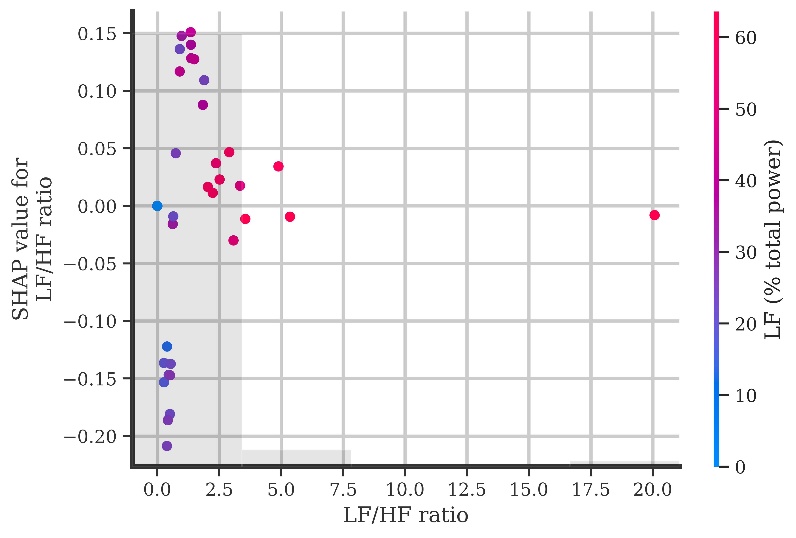

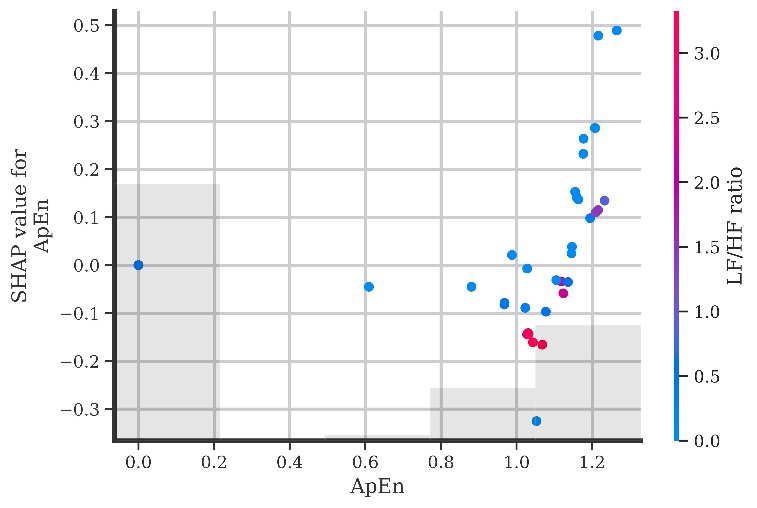

**Supplemental Figure 1.** SHapley Additive exPlanations (SHAP) dependence plots for the top three features identified from the pooled SHAP values, illustrating distinct patterns of feature influence on model predictions: relative low-frequency power as a percentage of total power (LF % total power) colored by approximate entropy (ApEn) **(top)**; the ratio of low-frequency to high-frequency power (LF/HF ratio) colored by LF % total power **(middle)**; ApEn colored by LF/HF ratio **(bottom)**.

**Supplemental Table 1.** Parameters and settings used for the machine learning pipeline. Abbreviations: traumatic brain injury (TBI); post-traumatic stress disorder (PTSD); heart rate variability (HRV); cross-validation (CV); receiver operating characteristic (ROC); area under the ROC curve (AUC); central processing unit (CPU).

| Category | Component | Parameter | Value / Description |
| --- | --- | --- | --- |
| Data Preparation | Class labels |  | TBI + PTSD = 1; TBI-alone = 0 |
|  | Feature matrix |  | Predefined HRV feature set |
|  | Outcome variable |  | Binary target variable |
| Missing Data Handling | Missingness assessment |  | Missingness assessed across all features |
|  | Imputation method |  | Median imputation |
|  | Application rule |  | Applied only if missing values were detected |
| CV | Outer CV | Type | Stratified k-fold |
|  |  | Number of folds | 5 |
|  |  | Data shuffling | Enabled |
|  |  | Random seed | 42 |
|  | Inner CV | Type | Repeated stratified k-fold |
|  |  | Number of folds | 3 |
|  |  | Number of Repetitions | 5 |
|  |  | Data shuffling | Enabled |
|  |  | Random seed | 42 |
| Feature Filtering | | Correlation metric | Spearman's rank correlation |
|  |  | Threshold | Absolute correlation > 0.90 |
|  |  | Matrix region | Upper triangular correlation matrix |
| Feature Selection | Selection algorithm | Method | Boruta |
|  | Base estimator | Model type | Random forest classifier |
|  |  | Number of trees | Auto |
|  |  | Maximum tree depth | 10 |
|  |  | Random seed | 42 |
|  |  | Parallel computation | All available CPU cores |
|  | Boruta configuration | Number of iterations | 50 |
|  |  | Tree count setting | Automatically determined |
|  |  | Significance threshold | α = 0.10 |
|  |  | Random seed | 42 |
|  | Feature retention |  | Confirmed and tentative features retained |
| Random Forest | Learning algorithm | Model | Random forest classifier |
|  | Reproducibility | Random seed | 42 |
|  | Parallel computation | Parallel computation | All available CPU cores |
|  | Hyperparameter optimization | Method | Exhaustive grid search |
|  |  | Scoring metric | ROC AUC |
|  |  | Validation scheme | Inner repeated stratified CV (3-fold, 5 repeats) |
|  | Hyperparameter grid | Number of trees | 100, 200 |
|  |  | Maximum tree depth | 3, 5 |
|  |  | Minimum samples required to split a node | 5, 10 |
|  |  | Minimum samples required in terminal nodes | 2, 5 |
|  |  | Feature sampling strategy | Square root of total features |
| Feature Scaling | Preprocessing | Scaling applied | None (tree-based models are scale-invariant) |
| Performance Metrics | Classification evaluation | Metrics computed | ROC AUC, accuracy, precision, recall, F1 score |
|  | Metric robustness | Zero-division handling | Undefined values set to zero |
|  | Class balance assessment | Additional metric | Balanced accuracy (computed from pooled predictions) |
| ROC Analysis | Curve construction | Method | Threshold-based ROC curve |
|  | Summary statistic | AUC computation | Trapezoidal integration |
